# The Verification Gap: Artificial Intelligence Adoption, Hallucination Awareness, and Verification Practices Among Early Career Medical Researchers in Pakistan

**DOI:** 10.64898/2026.05.28.26354373

**Authors:** Mobeen Sajjad

## Abstract

**Background:** Artificial intelligence (AI) tools have been rapidly adopted by medical researchers, yet whether early career researchers in low- and middle-income countries possess the awareness and habits needed to use these tools safely remains poorly documented. This study characterized AI adoption, hallucination awareness, and verification and disclosure practices among early career medical researchers in Pakistan.

**Methods:** A cross-sectional anonymous online survey was conducted among medical students, house officers, residents, physicians, and faculty involved in research or academic work across Pakistan (May 2026). Descriptive statistics and chi-square tests were applied to 373 eligible responses.

**Results:** AI use was near-universal (99.7%), with 60.3% using AI daily. The most commonly reported tool was Claude (40.5%), followed by ChatGPT (29.2%) and Perplexity (26.0%), though this ranking likely reflects sampling characteristics. Despite high adoption, 59.2% typically did not verify AI outputs before use, and 40.2% had never heard that AI can generate fabricated references. In behavioral vignettes, 36.5% assumed convincing AI-generated references were authentic, and 54.2% would continue using remaining AI content after discovering one fabricated reference. Formal research training was strongly associated with consistent disclosure (51.7% vs. 17.1%; chi-square=48.43, p<0.001). Role, daily use frequency, and research training were not significantly associated with verification behavior.

**Conclusions:** Early career medical researchers in Pakistan demonstrate high AI adoption alongside incomplete hallucination awareness and infrequent verification — a pattern that may carry implications for research integrity. Formal training was the only factor significantly associated with consistent disclosure. Integration of AI literacy into medical curricula and institutional governance frameworks merits consideration.

## 1. Introduction

The integration of artificial intelligence (AI) tools — particularly large language models (LLMs) such as ChatGPT, Claude, and Perplexity — into medical research workflows has accelerated considerably in recent years [1,2]. These tools offer practical advantages for researchers working under resource constraints: they can assist with literature synthesis, manuscript drafting, language editing, data interpretation, and reference generation at a speed and scale previously unavailable to independent investigators in low- and middle-income country (LMIC) settings [3,4].

However, a recognized limitation of contemporary LLMs is their tendency to generate content that appears scientifically credible but is factually incorrect — a phenomenon commonly referred to as hallucination [5,6]. Of particular relevance to research integrity is the generation of fabricated references: citations that appear authentic in format, authorship, and journal attribution, but do not correspond to real publications [7]. When incorporated into submitted manuscripts without verification, such fabricated references may compromise the reliability of the scientific record [8].

Existing literature on AI use in medical research has predominantly focused on adoption rates, perceived utility, and ethical attitudes [1,2,9]. Systematic documentation of verification practices and hallucination awareness remains limited, particularly in LMIC contexts [9]. Pakistan presents an important setting: it maintains one of the largest medical education systems in South Asia, has experienced notable growth in research output, and yet lacks formal AI governance frameworks or structured AI literacy training in most medical institutions [10].

This study was designed to address this gap. A cross-sectional survey characterized AI adoption patterns, hallucination awareness, and verification and disclosure practices among early career medical researchers across Pakistan. We hypothesized that AI adoption would substantially outpace the development of responsible verification and disclosure practices — a pattern we term the verification gap.

## 2. Methods

### 2.1 Study Design

We conducted a cross-sectional, self-administered, anonymous online survey, reported in accordance with the Strengthening the Reporting of Observational Studies in Epidemiology (STROBE) guidelines.

### 2.2 Setting and Participants

Participants were recruited from across Pakistan via WhatsApp medical research networks, LinkedIn, and direct professional contacts during May 2026. Eligible participants were individuals currently involved in medical education, clinical practice, research, or academic work in Pakistan, aged 18 years or older, who provided voluntary informed consent.

### 2.3 Survey Instrument

The survey was administered using Google Forms and comprised 24 items across four domains: (1) demographic and professional characteristics; (2) AI adoption patterns; (3) verification practices and hallucination awareness, including two behavioral vignettes presenting realistic AI use scenarios; and (4) attitudes and disclosure practices. Three items — gender, purpose of AI use, and disclosure practice — were added following initial piloting after 24 responses had been collected. These respondents were individually contacted and their answers incorporated into the final dataset. All responses were anonymous; no personal identifiers were collected.

### 2.4 Ethical Considerations

This study involved anonymous, voluntary participation with no collection of personal identifiers, no clinical intervention, and no patient data. All participants provided explicit informed consent. Under these conditions, the study qualifies for exemption from formal institutional review board review in accordance with standard criteria for anonymous survey research. The author declares that AI assistance in preparing this manuscript was limited to language editing; all content was independently reviewed and verified, and all references were manually confirmed. AI use is disclosed in accordance with journal policy.

### 2.5 Statistical Analysis

Data were analyzed using Python (pandas and scipy libraries). Frequencies and percentages were calculated for all categorical variables. Chi-square tests of independence assessed associations between demographic and training variables and key outcomes including verification behavior, hallucination awareness, and disclosure practice. A p-value of less than 0.05 was considered statistically significant. No corrections for multiple comparisons were applied given the exploratory nature of this study; results should be interpreted as hypothesis-generating rather than confirmatory.

## 3. Results

### 3.1 Participant Characteristics

A total of 373 individuals completed the survey, all confirming active involvement in medical education, clinical practice, research, or academic work in Pakistan. No responses were excluded. Participant characteristics are summarized in Table 1.

**Table 1.** Demographic and professional characteristics (N=373)

| Characteristic | N | % |
| --- | --- | --- |
| Current role: Medical student | 99 | 26.5 |
| Current role: Resident | 106 | 28.4 |
| Current role: House officer | 62 | 16.6 |
| Current role: Physician | 63 | 16.9 |
| Current role: Faculty | 42 | 11.3 |
| Gender: Female | 186 | 49.9 |
| Gender: Male | 185 | 49.6 |
| Publications: 0 | 1 | 0.3 |
| Publications: 1–2 | 240 | 64.3 |
| Publications: 3–5 | 93 | 24.9 |
| Publications: >5 | 38 | 10.2 |
| Formal research training: Yes | 151 | 40.5 |
| Formal research training: No | 219 | 58.7 |
| Previously participated in research: Yes | 292 | 78.3 |

The most common roles were resident (28.4%, n=106) and medical student (26.5%, n=99), followed by physician (16.9%, n=63), house officer (16.6%, n=62), and faculty (11.3%, n=42). Gender distribution was approximately equal (female 49.9%, male 49.6%). Responses were received from nine cities, with the largest proportions from Gujranwala (19.6%), Lahore (17.7%), and Faisalabad (13.4%). The majority (64.3%, n=240) reported 1–2 publications; 24.9% reported 3–5, and 10.2% reported more than five. Formal research methodology training had been received by 40.5% (n=151); 58.7% (n=219) reported no such training.

### 3.2 AI Adoption Patterns

AI use was near-universal: 99.7% reported using at least one AI tool. Daily use was reported by 60.3% (n=225), weekly by 25.5% (n=95), and monthly by 12.1% (n=45). The most commonly reported tool in this sample was Claude (40.5%, n=151), followed by ChatGPT (29.2%, n=109) and Perplexity (26.0%, n=97). This ranking likely reflects characteristics of the recruitment network and should not be interpreted as reflecting national AI adoption patterns. Common purposes for AI use included literature search, generating or finding references, interpreting data, and writing and editing manuscripts.

### 3.3 Verification Practices

Despite near-universal adoption, independent verification of AI outputs before use was uncommon. The majority (59.2%, n=221) reported typically not verifying outputs; only 1.3% (n=5) verified nearly all AI-generated content before use. A further 7.2% (n=27) reported trusting most AI outputs if they appeared scientifically convincing (Table 2).

**Table 2.**
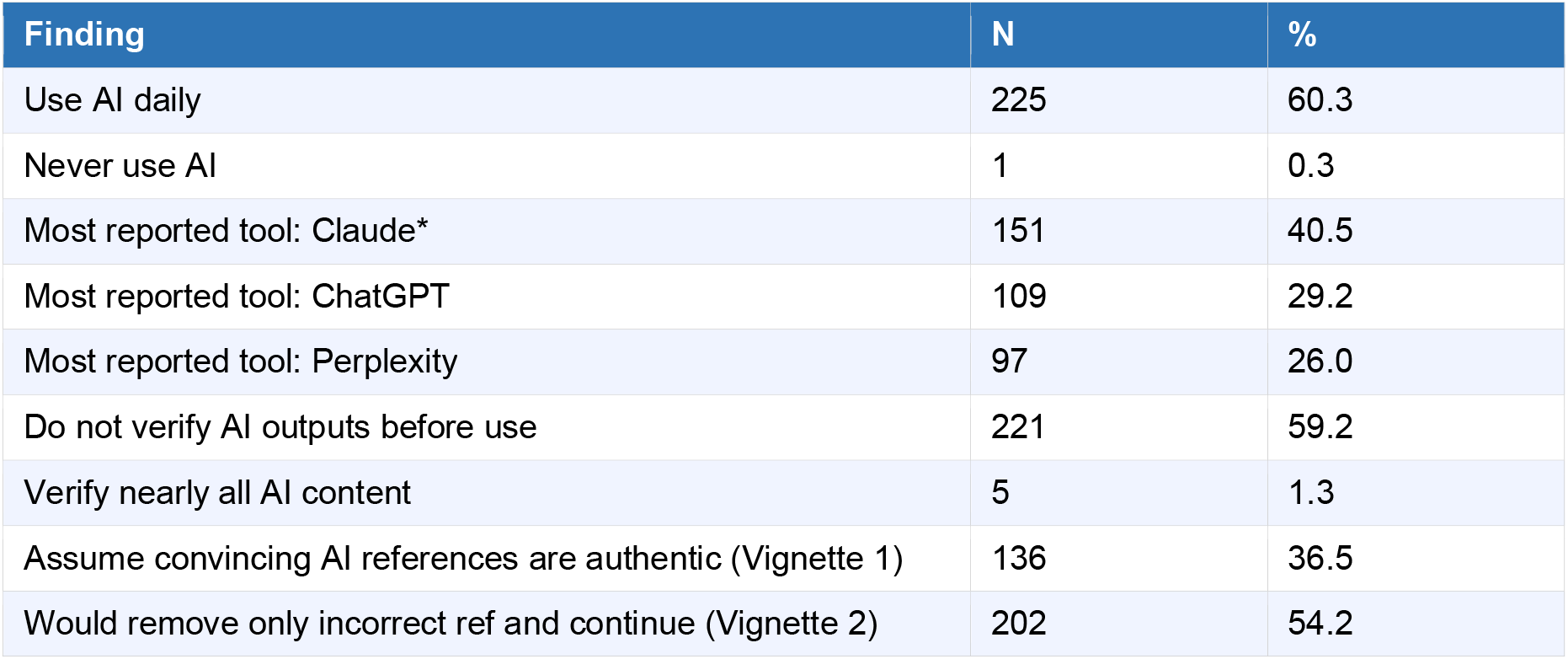
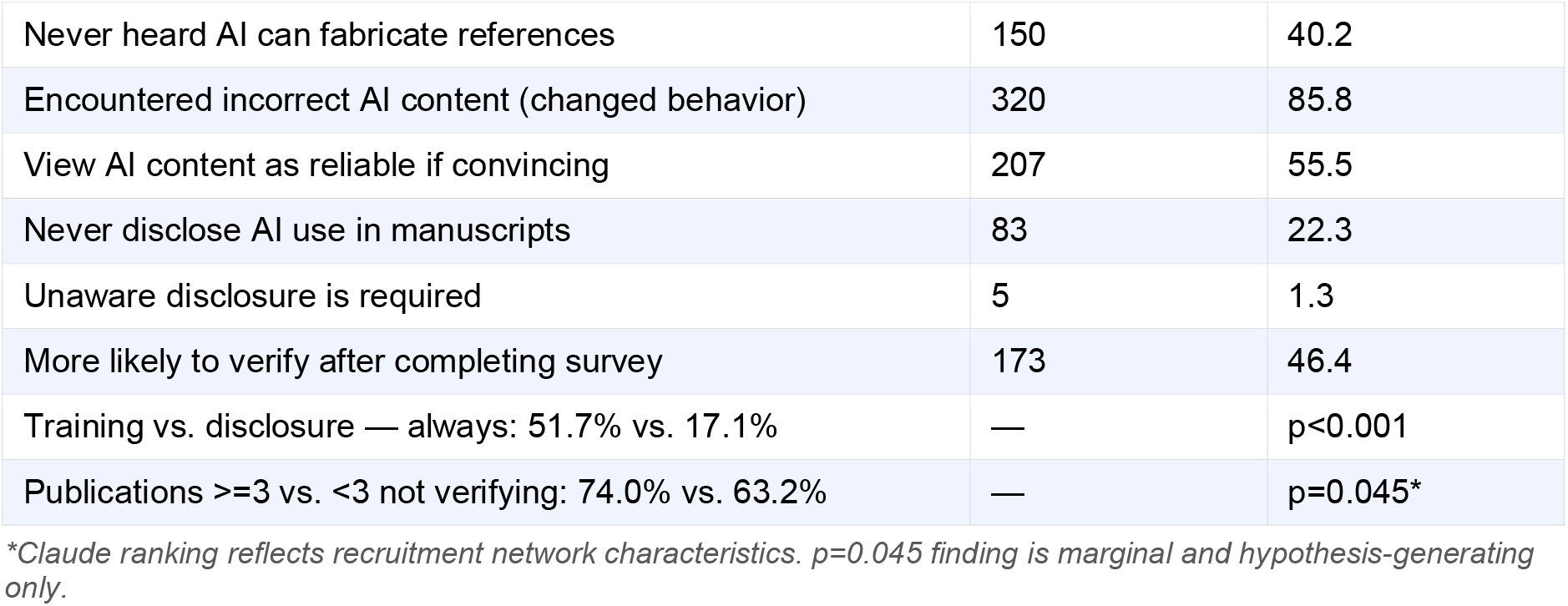
Key findings: AI adoption, verification, hallucination awareness, and disclosure (N=373)

In a behavioral vignette presenting AI-generated text with convincing-appearing references, 36.5% (n=136) assumed authenticity based on appearance alone, while 35.1% (n=131) recognized the need for independent verification. In a second vignette, 54.2% (n=202) would remove only the discovered fabricated citation and continue using remaining AI output, and 16.9% (n=63) would continue because occasional errors are expected. When asked about the speed-versus-reliability tradeoff, 49.6% (n=185) would prioritize speed even if some risk remained; 21.2% (n=79) would prioritize reliability even at the cost of efficiency.

Chi-square analysis revealed no statistically significant association between professional role and verification behavior (chi-square=0.81, df=2, p=0.67), daily versus non-daily use frequency (chi-square=0.09, df=1, p=0.77), or formal research training (chi-square=0.15, df=1, p=0.70). Researchers with three or more publications were somewhat less likely to report verifying outputs than those with fewer (74.0% vs. 63.2% not verifying; chi-square=4.03, df=1, p=0.045); however, given the marginal p-value and multiple comparisons conducted without correction, this finding should be considered hypothesis-generating only.

### 3.4 Hallucination Awareness

Awareness of AI hallucination was substantially incomplete. Among respondents, 40.2% (n=150) reported regularly using AI but having never heard that AI may generate fabricated scientific references; 27.9% (n=104) knew AI could make mistakes but had not considered fabricated references specifically; and 31.1% (n=116) reported prior full awareness. No significant association was found between professional role and awareness (chi-square=0.92, df=2, p=0.63), or between daily AI use and awareness (chi-square=0.00, df=1, p=1.00), suggesting this knowledge gap was broadly distributed regardless of seniority or intensity of use.

Despite limited conceptual awareness, 85.8% (n=320) reported personally encountering AI-generated information that proved incorrect or fabricated after verification, with this experience changing how they used AI. A further 8.0% (n=30) reported the same encounter but continued using AI in the same way.

### 3.5 Attitudes and Disclosure Practices

Over half of respondents (55.5%, n=207) viewed AI-generated content as generally reliable when it appeared convincing and included references; 16.1% (n=60) felt AI outputs should never be trusted without verification. Regarding disclosure, 22.3% (n=83) reported never disclosing AI assistance in manuscripts, and 1.3% (n=5) were unaware that disclosure is required. Only 31.1% (n=116) reported always disclosing; 44.8% (n=167) disclosed only sometimes.

A statistically significant association was observed between formal research methodology training and consistent AI disclosure: 51.7% of trained researchers reported always disclosing, compared with 17.1% of untrained researchers (chi-square=48.43, df=1, p<0.001). This was the strongest and most robust association identified in this dataset.

### 3.6 Survey-Related Attitude Change

When asked whether completing the survey changed their perspective on verifying AI-generated content, 46.4% (n=173) reported being more likely to verify outputs following participation; 17.4% (n=65) reported already routinely verifying. These findings suggest that brief structured exposure to hallucination concepts may influence verification attitudes, though the absence of a longitudinal design precludes conclusions about the durability or causal nature of this effect.

## 4. Discussion

This cross-sectional survey of 373 early career medical researchers across Pakistan suggests a meaningful discrepancy between the intensity of AI adoption and the verification and disclosure practices needed to support its responsible use in academic research. AI use was near-universal and daily, yet hallucination awareness was substantially incomplete, independent verification was infrequent, and AI use was regularly undisclosed in manuscripts. These patterns were broadly consistent across professional roles and AI use frequency; formal research methodology training was the only factor significantly associated with improved disclosure behavior.

The high prevalence of AI use — 60.3% reporting daily use — is consistent with international data documenting rapid integration of AI into medical research workflows [1,2]. Against this background, the finding that 40.2% of respondents had never encountered the concept of AI-generated fabricated references raises a practical concern: limited awareness of hallucination risk may reduce the likelihood of systematic verification among researchers who might otherwise verify if they understood the risk. This inference remains speculative given the cross-sectional and self-report nature of the data; actual verification behavior and its relationship to awareness would require prospective or observational methods to confirm.

The behavioral vignette data offer additional perspective. When presented with convincing-appearing AI-generated references, fewer than four in ten respondents recognized the need for independent verification. When confronted with a discovered fabricated reference, 54.2% would remove only that citation and continue using surrounding AI output — a response that reflects incomplete understanding that AI errors may affect any part of generated content, not only the specific citation identified. These responses are consistent with the broader self-reported verification patterns and together support the hypothesis that adoption has outpaced the development of critical appraisal habits.

The marginally significant association between publication volume and verification behavior (p=0.045) warrants cautious interpretation. Researchers with three or more publications were somewhat less likely to verify AI outputs, which could reflect automation bias — the tendency to over-rely on automated tools as they become embedded in habitual workflows [12]. However, given the multiple comparisons conducted without correction and the borderline p-value, this finding should be treated as hypothesis-generating and not cited as a definitive result.

The most robust finding in the inferential analysis was the strong association between formal research training and consistent AI disclosure (51.7% vs. 17.1%, p<0.001). This suggests that structured exposure to research norms — including ethics, transparency, and publication standards — may support responsible AI use practices. Importantly, training was not significantly associated with verification behavior, pointing to a dissociation between normative awareness (knowing disclosure is required) and habitual practice (actually verifying content). This distinction may have practical implications for how AI literacy interventions are designed.

Framing these findings within their systemic context is important. The observed discrepancy between adoption and responsible practice reflects not primarily a reflection of individual negligence but the absence of institutional infrastructure: no standardized AI literacy curriculum in Pakistani medical education, no institutional verification protocols, and limited enforcement of disclosure norms. Structural interventions — integration of AI literacy training into medical curricula and establishment of institutional AI governance frameworks — are therefore more likely to be effective than those targeting individual behavior alone.

The apparent influence of survey participation on verification attitudes — with 46.4% reporting increased likelihood of verifying outputs after completing the survey — suggests that brief structured exposure to hallucination concepts may be sufficient to shift attitudes. This finding is preliminary and should not be overinterpreted; the absence of a control condition and longitudinal follow-up means the durability and causal nature of this effect cannot be established from these data.

Several limitations should be acknowledged. Convenience sampling via WhatsApp and LinkedIn introduces selection bias; respondents may be more research-active or digitally engaged than the broader population of early career Pakistani medical professionals. Findings may not generalize to non-English-speaking or non-digitally engaged healthcare professionals. Self-reported data are subject to social desirability bias, potentially overestimating verification and disclosure practices. The cross-sectional design precludes causal inference. The ranking of Claude as the most commonly reported tool likely reflects investigator network characteristics and should not be generalized to national patterns. Three questions were added after initial data collection, with missing responses subsequently obtained manually. Finally, this study measured self-reported perceptions and behaviors, not directly observed research conduct; actual rates of verification and fabricated reference use may differ from those reported here.

## 5. Conclusions

This cross-sectional survey suggests that early career medical researchers in Pakistan have adopted AI tools at a rate that substantially outpaces the development of verification habits, hallucination awareness, and disclosure practices. Formal research methodology training was significantly associated with consistent AI disclosure, though not with verification behavior — a dissociation that may inform the design of future educational interventions. These findings provide a quantitative basis for advocating the integration of AI literacy, including explicit training on hallucination risk and verification practice, into Pakistani medical education curricula and institutional research governance frameworks. Replication using probability-based sampling across broader LMIC contexts is warranted.

## Declarations

### Author Contributions

Mobeen Sajjad: conceptualization, study design, data collection, data analysis, manuscript writing, and final approval.

### Data Availability

The anonymized dataset will be made available upon reasonable request to the corresponding author.

### AI Use Disclosure

AI tools were used for language refinement during manuscript preparation. All scientific content, interpretations, and references were developed and independently verified by the author.

## STROBE Compliance Statement

This manuscript was prepared in accordance with the STROBE statement for cross-sectional studies. A completed STROBE checklist is available upon request from the corresponding author.

